# Will there be a third COVID-19 wave? A SVEIRD model based study of India’s situation

**DOI:** 10.1101/2021.05.16.21257300

**Authors:** Dwarakesh Kannan, R Gurusriram, Rudra Banerjee, Srijit Bhattacharjee, Pritish Kumar Varadwaj

## Abstract

Since first patient detected in India in late February, 2020, SARS-CoV-II virus is playing havoc on India. After the first wave, India is now riding the 2nd wave. As was in the case of European countries like Italy and UK, the 2nd wave is more contagious and at the time of writing this paper, the per day infection is as high as 400,000. The alarming thing is it is not uncommon that people is getting infected multiple time. On the other hand, mass vaccination has started step by step. There is also growing danger of potential 3rd wave is unavoidable, which can even infect kids and minors.

In this situation, an estimation of the dynamics of SARS-CoV-2 is absolutely necessary to combat the pandemic. We have used a modified SEIRD model, that includes vaccination and repeat infection as well. We have studied India and 8 Indian states with varying SARS-CoV-2 infection. We have shown that, Covid-19 wave will be repeated time to time, but the intensity will slow down with time. In most possible situation, our calculation shows COVID-19 will remain as endemic for foreseeable future, unless we are able to increase our vaccination rate manifold.

## 1 Introduction

COVID-19 was announced as a global pandemic by World Health Organization (WHO)[1] owing to its highly contagious and pathogenicity that has been rapidly spreading throughout the world since its first reported outbreak in China in December, 2019. The “first wave” of COVID-19 slowed down by September-October,2020 as was correctly predicted by three of the current authors[2]. After going down to as low as 10000 active patient, the “second wave” picked up by the middle of March, 2021 in India. Typical to the 2nd wave seen in European countries, this is more infectious and hence more fatal[3], as it has pushed the health-infrastructure of the country to its limit. As a ray of hope, the vaccination programme is going on full force throughout the country[4]. On the other hand, repeat infection of COVID-19 is not uncommon either.

As it is already noted, the 2nd wave has pushed our health infrastructure to its limit. To overcome this situation, administrations and health departments pan India, with 1.3 billion population and average population density of 382 persons per km^2^ need a very clear and through idea of the challenges they are facing in this time of pandemic. An early estimation of the width of the spread and the peak will give a front line fighters an idea what they need to do. This will also enable the government to make a decisive planning and management of resources efficiently. Hence a detailed analysis of real data is very much helpful to estimate possible distribution of testing kits, oxygen, medicines, hospitals, isolation centers etc.

The motivation of present work is to forecast the pandemic with the help of SVEIRD model, a well studied model for studying the dynamics of infectious diseases. With more than 1 years data in deposition, both in government and public sources, we can nowcast and forecast the dynamics of COVID-19. We have assessed different states with different population and infection density to show the dynamics. We have chosen 3 states with high populations density (Kerala, Tamil Nadu and Uttar Pradesh) with population density(*pd >* 500*/km*^2^), 3 states(Goa, Gujarat and Karnatak) with medium population density (300*/km*^2^ *< pd <* 500*/km*^2^) and 2 states (Chhattisgarh and Madhya Pradesh) with less population density (*pd <* 300*/km*^2^). Each state in these population density region is further catagorized by infection density (id, percentage of infected population). We have taken *id >* 2%, 1% *< id <* 2% and 1% *> id* as high, medium and low infection.

Rajesh Ranjen *et al*.[5] has recently characterised the parameters of COVID-19 based on total positivity rate, case fatality and effective reproductive rate. They have used SIR model for the prediction. This, together with work on testing reported by Cherian *et al*.[6] with more complicated version of SIRD model shows the necessity of more testing and a dependable source for people hospitalized or in home isolation. In a new model based analyses, including asymptomatic and undetected population, Agarwal et al. [7] have showed the appearance of peak for the second wave in different states of India quite accurately. In this paper, with a simpler model and a different approach we have analyzed the same. Moreover the possible occurrence of third wave is also predicted.

Overall, while the largest vaccination drive is going on in the history of human civilisation[8] in the country, it is crippled by slow vaccination rate. In the same time, government is talking about the third wave while the second wave is far from over. In this situation, COVID-19 infection dynamics would lead us for better preparation and hopefully save more life, absence of which in 2nd wave lead to the current chaos and death[9].

## 2 Methodology

We have employed a modified version of the standard epidemiological model for infectious disease, i.e. SEIRD model[2] to include effect of repeated infection and vaccination. Let us describe the effect briefly.

### 2.1 Data source

The data for daily number of people infected, recovered and deceased from 1st January to 30th April, 2021 are collected from publicly available database like covid19india.org[10] and worldometer[11]. For overall scenario, Indian government’s websites[12, 13] are consulted. From these available data, which are in comma separated value(csv), a few pre-processing were carried out to feed the SVEIRD based model calculations, which is written in python-3. The population density was collected from 2011 census[14].

### 2.2 SVEIRD Model

To analyze the data, we have implemented Susceptible(S)-Vaccinated(V)-Exposed(E)-Infectious(I)-Recovered(R)-Dead(D) model. In the latent phase of COVID-19 infection, the infected individual is not infectious. This latent period can be incorporated in simpler SIR model by adding the latent/exposed population (E). The infected population moves from S to E and from E to I and hopefully to R(Figure (1)). In this model, we have considered vaccinated people is not susceptible. This is parameterized in absence of real data.

**Figure 1:**
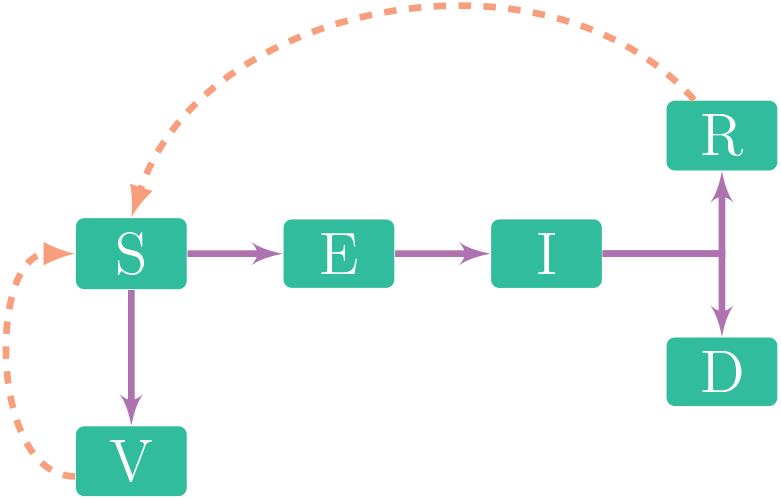
Dynamics of SVEIRD model

The dynamical equations for SVEIRD model are:

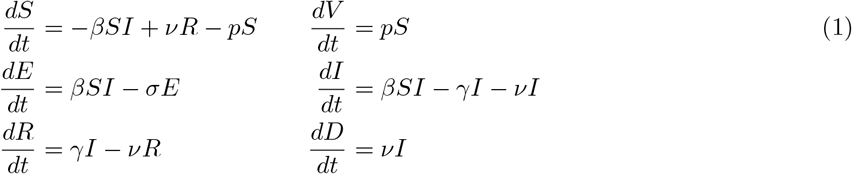

with initial conditions *S*(0) *>* 0, *E*(0) ≥ 0, *I*(0) ≥ 0, *V* (0) ≥ 0 and *R*(0) ≥ 0. Constants *β, p, γ, σ, ν* are defined in section(2). Mortality rate can be calculated from Covid-India data and is related to *ν*. The recovery rate is inverse of mean infectiousness period, that is average time to get cured from the onset of symptoms. The recovery rate is taken as 3, 4 and 5 days, as suggested by Bi *et al*. [15] from the studies based on Wuhan, inverse of which gives *γ* in Equation (1).

### 2.3 Descriptions of the coefficients

Let us now describe the definitions and method of computation of the parameters for studying the population dynamics with the help of SVEIRD model. Since the second wave has started at different time in different states, we have taken the starting point of our calculation from January 1, 2021 (120 days).

#### 2.3.1 Basic Reproductive Ratio(*R*_0_)

The reproduction rate *R*_0_ can be calculated with the daily cases starting from day 1 (January 1, 2021). Assuming that daily incidence obeys an approximate Poisson distribution, one employs Maximum Likelihood Estimate method. Given observation of (*N*_0_, *N*_1_, …, *N*) incident cases over consecutive time units, and a generation time distribution *w, R*_0_ is estimated by maximizing the log-likelihood

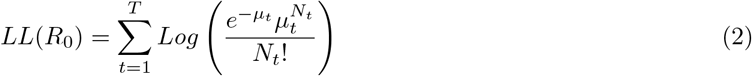

where 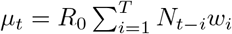. The random vector *w*_*i*_, can be estimated as mean doubling time (Figure (12b)), mean time taken to have number of cases to become double in a given period. *w* is also called serial interval distribution [16, 17].

*R*_0_ calculated for data upto 30th April, 2021 produced a value between 1.5 *−* 4 [18]. The value of *R*_0_ for India is estimated to be around 2.044 with the data up to this time limit.

#### 2.3.2 Serial Interval(*T*_*g*_)

The serial interval is the sum of incubation period (*d*_*ic*_) and infectious period(*d*_*if*_). Incubation period is the lag in days between being infected and developing the symptoms. Whereas infectious period is defined as the number of days an infected person remains infectious or can infect another person, after getting infected. Doubling time is defined as time period in which the cumulative infection doubles [19, 20]. Doubling Time is a function of both *T*_*g*_ and *R*_0_[21].

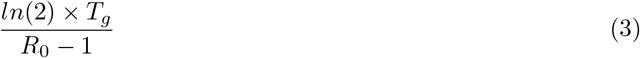

#### 2.3.3 Recovery Rate(*γ*)

The recovery rate is defined as the inverse of infectious period *d*_*if*_. The data from WHO and other studies[22, 23, 24, 15] show the infectious period is between 3-5 days.

#### 2.3.4 Incubation rate(*σ*)

Incubation rate is calculated by taking inverse of mean incubation period *d*_*i*_*n*. The data from WHO and other studies[25, 16, 26, 23, 24, 15] show the incubation period is between 5-7 days.

#### 2.3.5 Mortality Rate(*ν*)

The mortality rate is calculated as number of individuals died on a given period of time divided by the total infected in that period [27].

#### 2.3.6 Vaccination Rate(*p*)

The vaccination rate indicates the rate at which vaccination has occurred in each states. This is calculated as number of people got their 2nd dose on or before 15th April, as immunity develops at least 2 weeks after the 2nd dose. We have also taken vaccination efficacy of 80%, as reported by Ministry of Health and Family welfare, Govt. of India[28]. Mathematically, this is expressed as

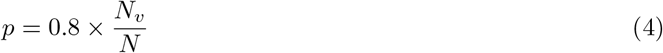

where *N*_*v*_ is the number of persons got 2nd vaccine till 15th April and *N* is the total population of the state.

## 3 Results

We applied the above SVEIRD model in 8 Indian states with varying population and SARS-CoV-2 infection density. Table (2a) gives the list of states chosen, with the population and infection density. The criteria of high, medium and low density is as described in section (1). In the table, high, medium and low is color-coded as red blue and green respectively. The same data is shown in Figure (2b) as a heatmap. We have calculated the SVEIRD dynamics of each of these states. From Figure (3) to Figure (11) we have shown the SVEIRD dynamics for infected population of selected states and of India as whole.

**Figure 2:**
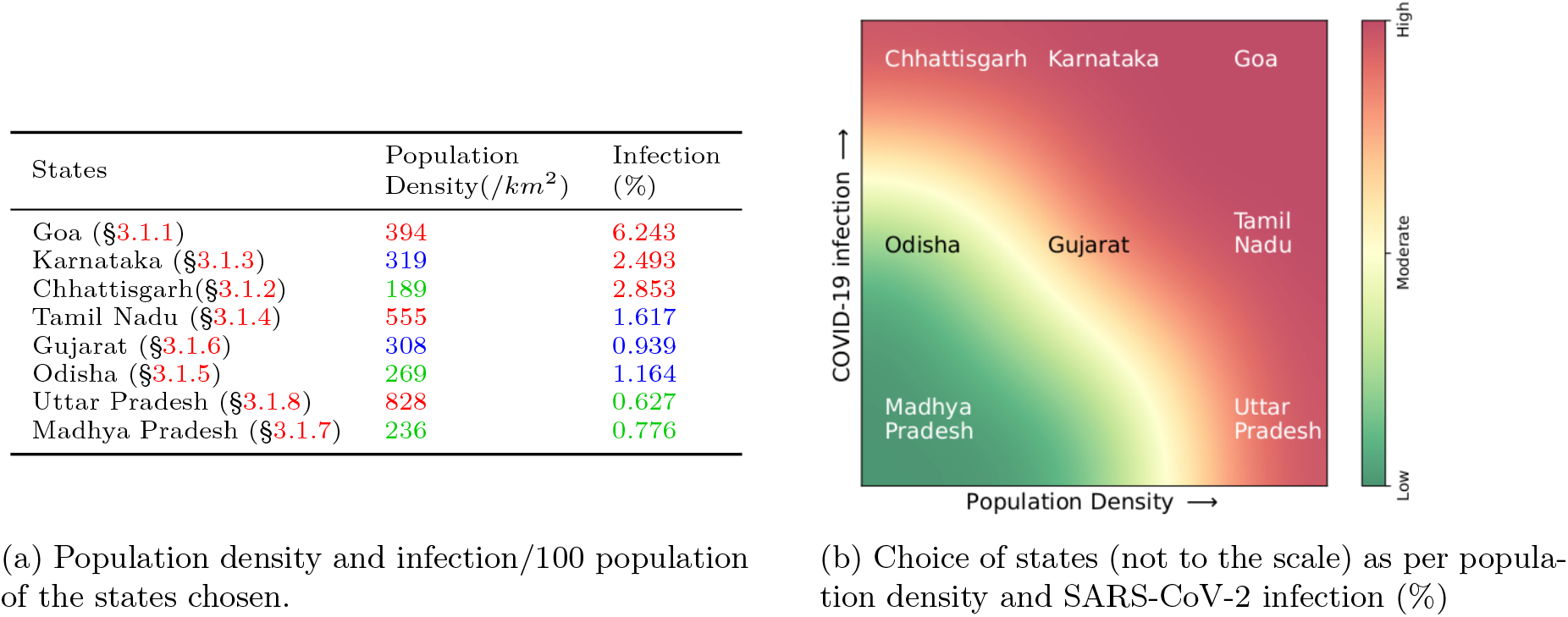
Choice of states and the SARS-CoV-2 infection % w.r.t. total population of the state and population density (/*km*^2^)

### 3.1 States

#### 3.1.1 Goa

a small south Indian state, has highest infection ratio in India(*≈*6%) and high population density(394 persons/*km*^2^) as shown in table (2a). According to our calculation, the *R*_0_ of Goa is 2.027. This also shows that this 2nd wave of COVID-19 in Goa will be over by May, at latest. The SVEIRD result for confirmed people is shown in Figure (3). Possibly due to their small population, Goa doesn’t show any sign of further wave in our range of study.

**Figure 3:**
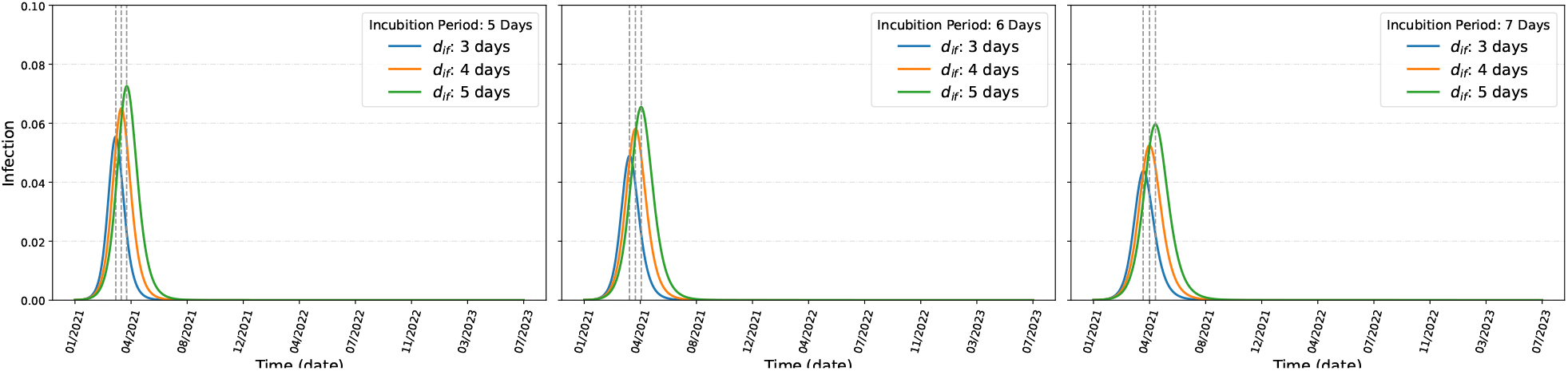
SVEIRD dynamics of Goa

#### 3.1.2 Chhattisgarh

is a medium size state in central India, with very less population density in Indian standard. It has population density 189 persons/*km*^2^ and high COVID-19 infection(2.853%) as shown in table (2a). As per our calculations, the *R*_0_ of Chhattisgarh is 1.810. Our calculation shows the peak of the second wave will arrive between end of April to end of September, as shown in Figure (4).

**Figure 4:**
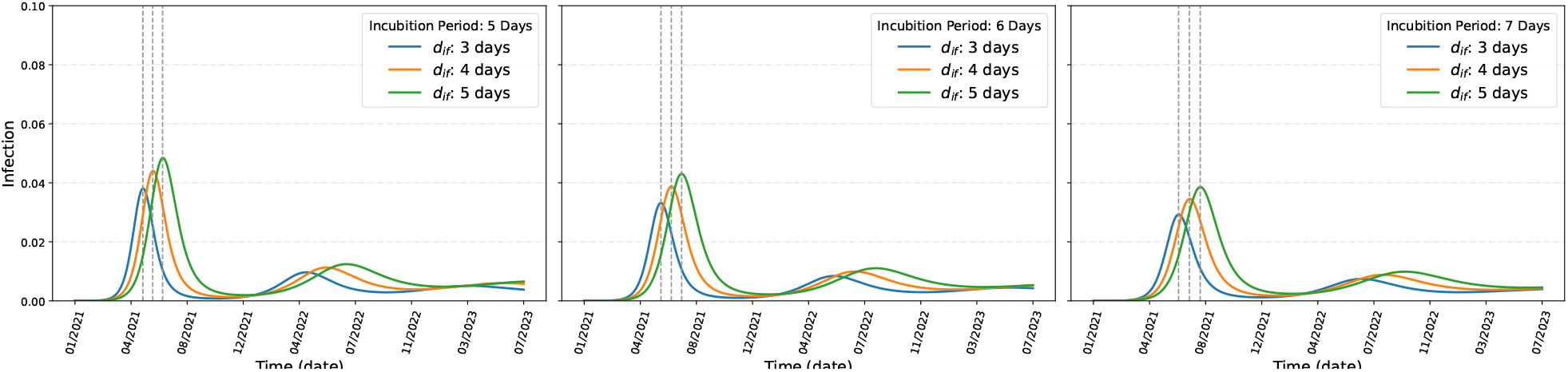
SVEIRD dynamics of Chhattisgarh

**Figure 5:**
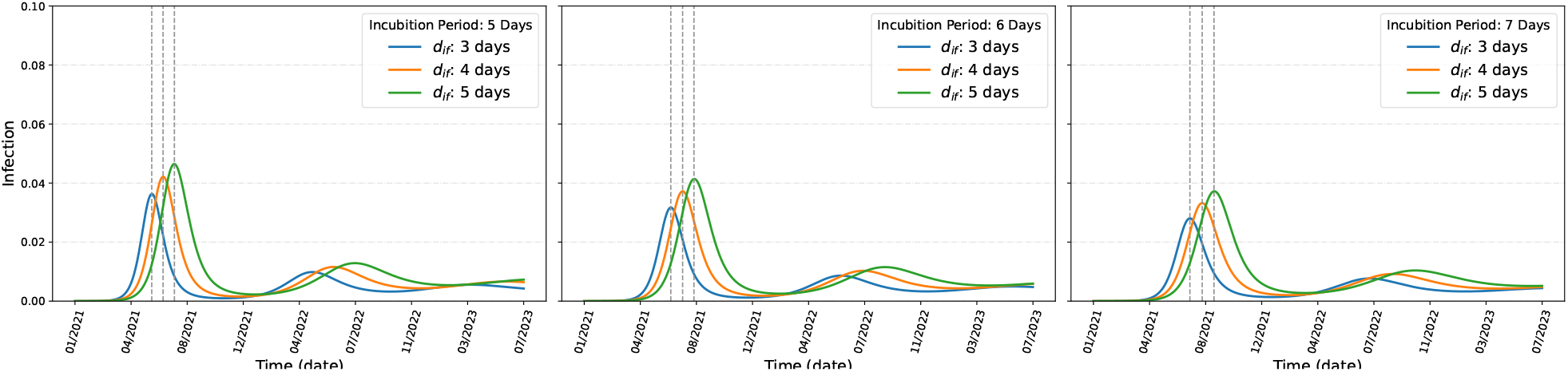
SVEIRD dynamics of Karnataka

**Figure 6:**
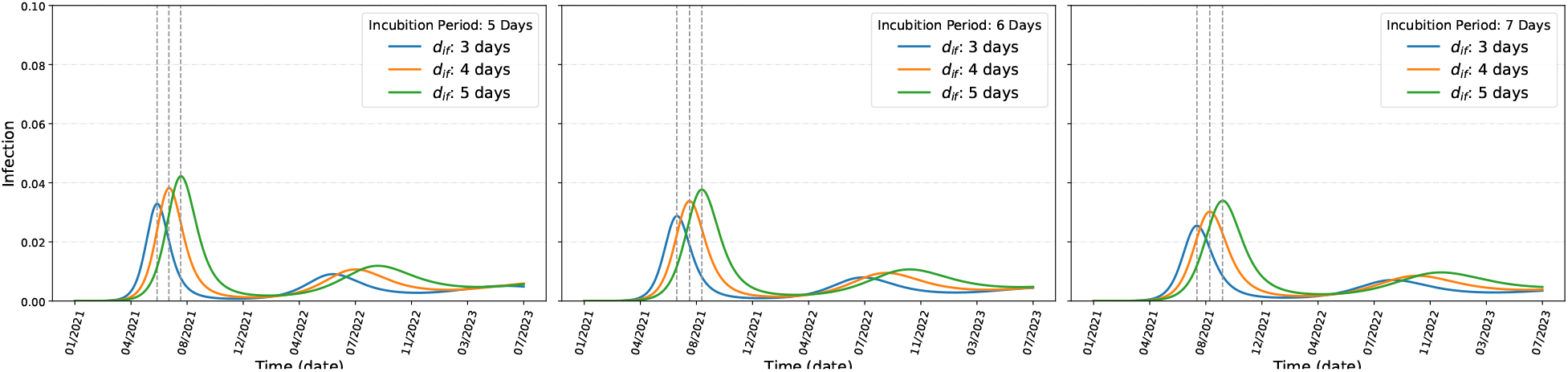
SVEIRD dynamics of Tamil Nadu

Though, the most dangerous feature is that, as per our calculation, the SARS-CoV-2 infection may not be completely over in Chhattisgarh with 2nd wave. A 3rd wave is very evident by middle of 2022. The data actually shows the COVID-19 may not be eradictated in Chhattisgarh and may live there as endemic.

#### 3.1.3 Karnataka

is one of the largest state in deccan peninsula, with medium (319 persons/*km*^2^) but high(2.853%) SARS-CoV-2 infection, as shown in table (2a). As of 30th April, 2021, Karnataka is already showing *≈*40,000 confirmed cases per day and there is no hint of slowing down. As per our calculations, *R*_0_ of Karnataka is 1.769. The peak of 2nd wave may happen between May to November. Just like Chhattisgarh, unfortunately, Karnataka also shows strong evidence of a 3rd wave by middle of next year.

#### 3.1.4 Tamil Nadu

is the southern most state of India, with high population density(555 persons/*km*^2^) but medium infection(1.617%), as shown in table (2a). Tamil Nadu shows dynamics of SARS-CoV-2 infection similar to its neighbour Karnataka. As per our calculations, the *R*_0_ of Tamil Nadu is 1.705. The SVEIRD calculation shows the peak of 2nd wave may be over between May to November, similar to that of Karnataka. Also, it shows that the 3rd wave is inevitable by the middle of next year. Unfortunately, Tamil Nadu also shows evidence that the pandemic will stay as an endemic in foreseeable future.

#### 3.1.5 Odisha

is a coastal state in eastern India, with low population density (269 persons/*km*^2^), as shown in table (2a). The *R*_0_ of Odisha is 2.245. Our study shows that Odisha has with all probability has crossed the peak of 2nd wave, which is between April and May, as shown in Figure (7). Also, our calculation shows very little sign of subsequent wave in the state of Odisha.

**Figure 7:**
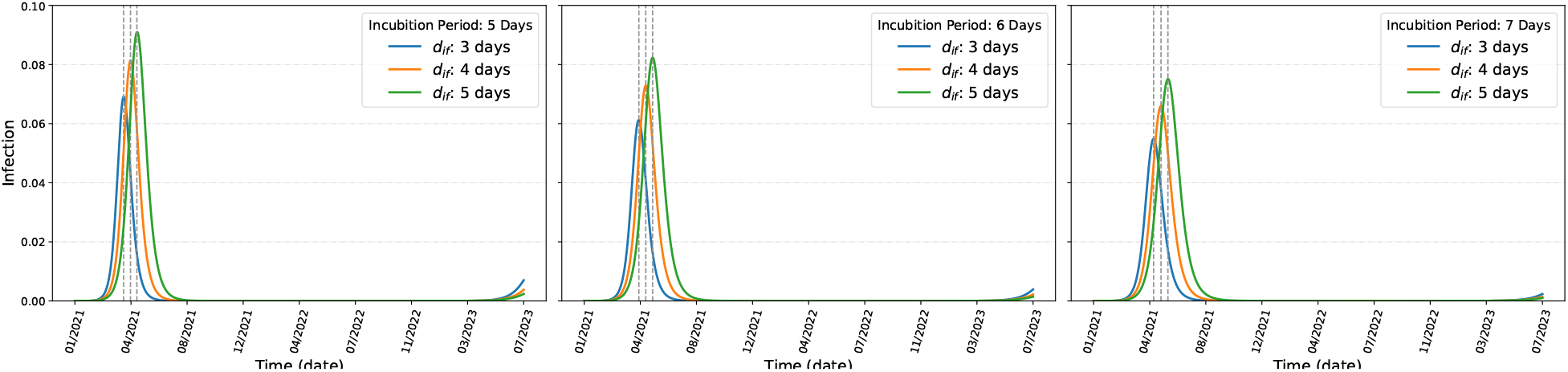
SVEIRD dynamics of Odisha

#### 3.1.6 Gujarat

is the west most state of India, with medium population density (308 persons/*km*^2^) and medium SARS-CoV-2 infection ratio (0.939%), as shown in table (2a). Our calculation shows *R*_0_ of Gujarat. The SVEIRD based dynamics shows the peak of 2nd wave will appear between May to July, as shown in

Figure (8). Gujarat also shows strong signal of subsequent 3rd wave and also that the COVID-19 will become an endemic in Gujarat.

**Figure 8:**
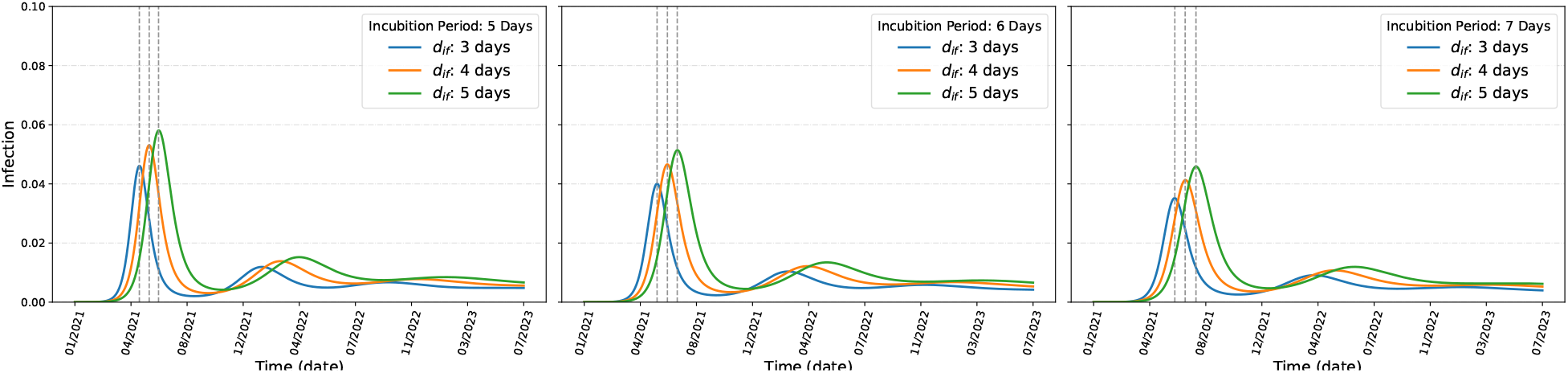
SVEIRD dynamics of Gujarat

#### 3.1.7 Madhya Pradesh

is the second largest state of India. It has a low (235 persons/*km*^2^) population density and low(0.776%) COVID-19 infection. The *R*_0_ of Madhya Pradesh is 2.054, as shown in table (2a). Our SVEIRD calculation shows the peak of the 2nd wave will be reached between April and September, as shown in Figure (9). Our calculation further shows that between 2nd and possible 3rd wave, Madhya Pradesh will be virtually COVID-19 free state. The infection will start again by the end of December, 2021 and will continue as endemic.

**Figure 9:**
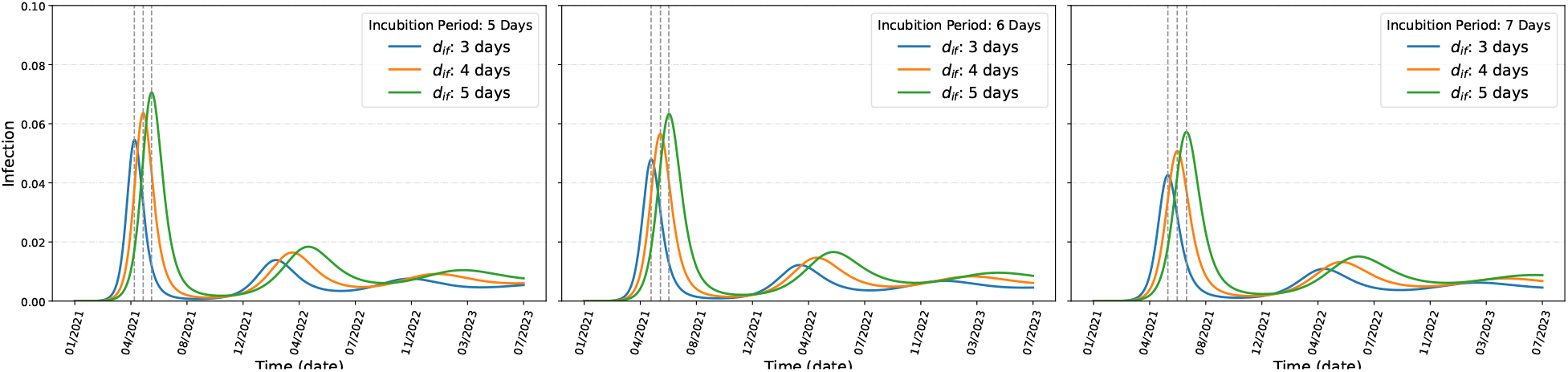
SVEIRD dynamics of Madhya Pradesh

#### 3.1.8 Uttar Pradesh

is the Indian state with highest population. It has a very high (828 persons/*km*^2^) population density with very low COVID-19(0.627%), as shown in table (2a). Our calculation shows the *R*_0_ of Uttar Pradesh is 3.217, which is highest among the states studied. The SVEIRD calculation shows the 2nd wave has crossed its peak on April, as shown in Figure (10). This matches the real data. This happy finding is overridden by the fact that Uttar Pradesh will be among the first to hit by the 3rd wave, which may start as soon as August, 2021. As normal to other populous states, SARS-CoV-2 will remain as endemic in Uttar Pradesh as well.

**Figure 10:**
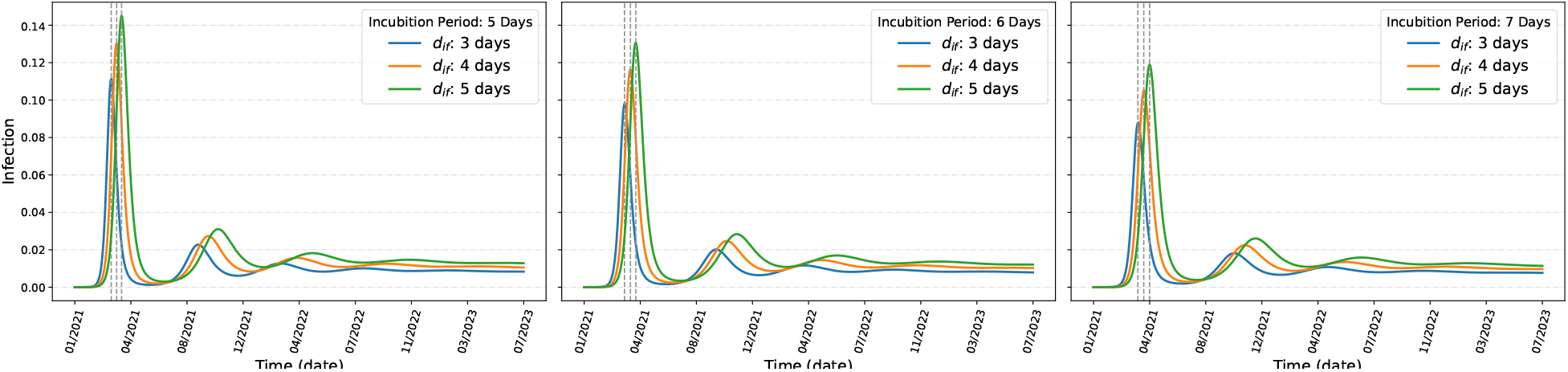
SVEIRD dynamics of Uttar Pradesh (Note the change of scale in *y-axis*)

### 3.2 India

Now, let us consider the status of India as a whole. The country with second highest population and very high population density, along with the unique socio-economic structure, it is almost impossible to maintain a social distance in India without a lockdown. This has lead to a catastrophe in India, with health system is running at its limit. Our calculation shows the *R*_0_ of India is 2.044. The SVEIRD calculation shows India will pass the peak of 2nd wave between June to October. The 3rd wave will start very soon after that, between December, 2021 to April, 2022, as evident from Figure (11).

**Figure 11:**
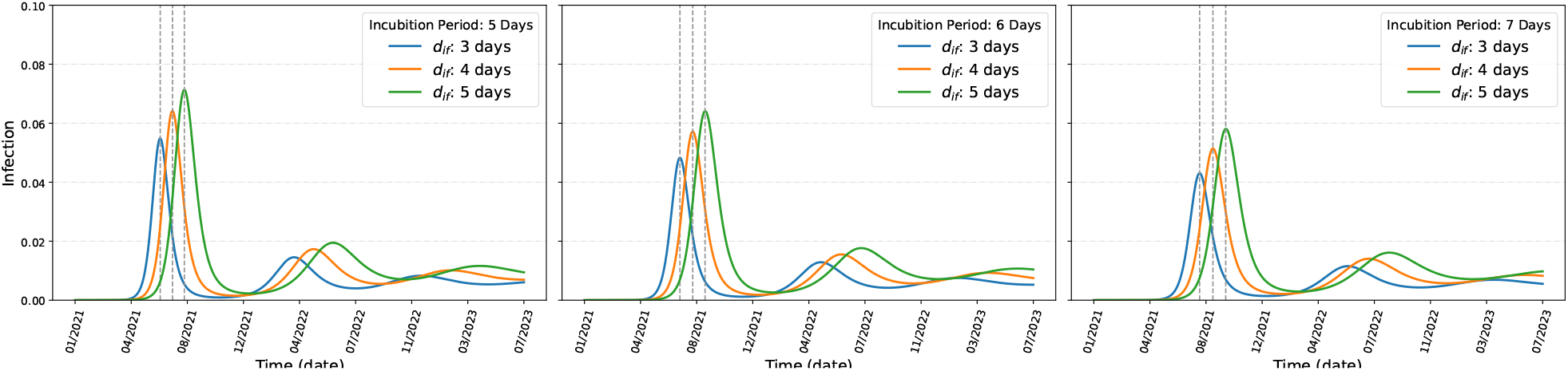
SVEIRD dynamics of India

A simple Gaussian fitting with data upto 30th April, 2021 shows the peak of 2nd wave will be around end of May(Figure (12a)), with per day infection could be as high as 700,000 persons. In the period of study, the mean doubling time is 20 days(Figure (12b)). This is consistent with the prediction of the Figure (12a) as the rate of infection on 30th April, 2021 is around 400,000.

**Figure 12:**
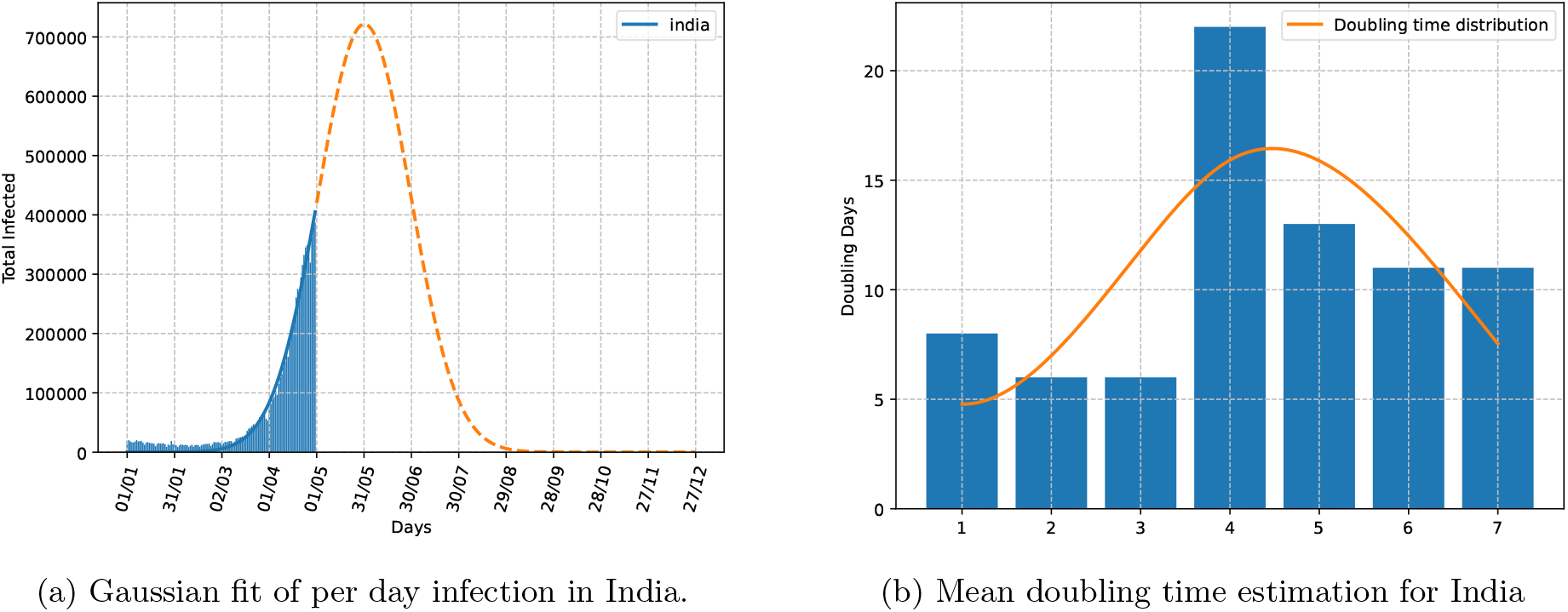
Variation of daily and cumulative incidence of COVID-19 cases in India

Finally, we have estimated the increase in the rate of vaccination we have to make to avoid the third wave. We have taken incubation period of 5 days and *d*_*if*_ of 5days for this study. Our calculations shows (Figure (13)) with increase in vaccination, the infection will slow down. To avoid 3rd wave, we have to increase the vaccination more the 5 times of the current vaccination rate.

**Figure 13:**
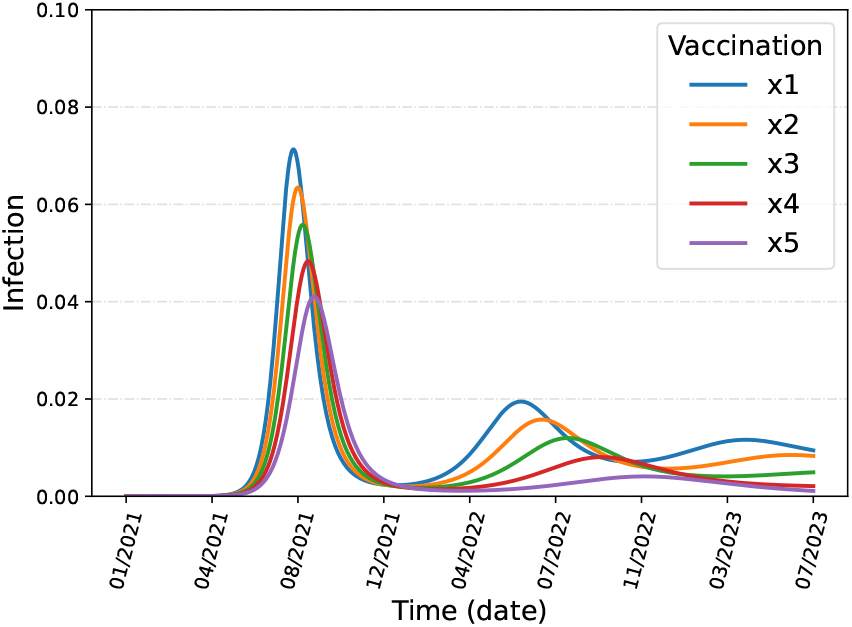
Change of SARS-CoV-2 infection with increase in vaccination rate in India. The line shows increase in rate of vaccination times the current rate. This is done for dynamics of India (Figure (11)) with incubation period 5 days and *d*_*if*_ 5 days.)

## 4 Conclusions

We have studied 8 states with varying population density and infection density pan India. Taken vaccination drive and repeat infection into consideration, we have shown the dynamics of SARS-CoV-2 infection. We have shown that the 3rd wave is inevitable and will be coming very soon. Though our dynamics shows, the 3rd wave will infect less number of people and more than 1 year away, we have to keep in mind that the SARS-CoV-2 virus is still involving and with highly contagious nature of the virus can make the 3rd wave come sooner and bigger. There is still no conclusive proof of longivity of the antibody created due to vaccine or infection. Until we get everyone vaccinated, a single superspreader may create catastrophe as is in the case of 2nd wave. Our study shows the mask and social distancing as a normal part of our daily life for foreseeable future.

## Data Availability

No human data used. Data source is cited properly.

## 5 Acknowledgements

DK, GR and RB acknowledge the infrastructural supports and High Performance Computing Center, SRM Institute of Science and Technology for providing the computational facility.

SB and PV acknowledge the infrastructural supports provided by IIIT Allahabad.

## References

[1] WHO director-general’s remarks at the media briefing on covid-19. https://www.who.int/dg/covid-19---11-march-2020, 2020. Accessed on 6th May, 2021.

[2] Rudra Banerjee, Srijit Bhattacharjee, and Pritish Kumar Varadwaj. Analyses and forecast for covid-19 epidemic in india. medRxiv, 2020.

[3] Andrea Borghesi, Salvatore Golemi, Nicola Carapella, Angelo Zigliani, Davide Farina, and Roberto Maroldi. Lombardy, northern italy: COVID-19 second wave less severe and deadly than the first? a preliminary investigation. Infectious Diseases, 53(5):370–375, feb 2021.

[4] Sanjeet Bagcchi. The world’s largest covid-19 vaccination campaign. The Lancet. Infectious Diseases, 21(3), 2021.

[5] Rajesh Ranjan, Aryan Sharma, and Mahendra K. Verma. Characterization of the second wave of covid-19 in india. medRxiv, 2021.

[6] Philip Cherian, Sandeep Krishna, and Gautam I. Menon. Optimizing testing for covid-19 in india. medRxiv, 2021.

[7] Manindra Agrawal, Madhuri Kanitkar, and Mathukumalli Vidyasagar. SUTRA: An Approach to Modelling Pandemics with Asymptomatic Patients, and Applications to COVID-19. arXiv e-prints, January 2021.

[8] 2021 by Ministry of Health Press Release on 16th January and Family Welfare. https://pib.gov.in/PressReleasePage.aspx?PRID=1689112. Accessed on 12th May, 2021.

[9] UNITED NATIONS DEVELOPMENT PROGRAMME:Human Development Reports. http://hdr.undp.org/en/content/global-preparedness-and-vulnerability-dashboards. Accessed on 12 May,2021.

[10] https://www.covid19india.org/.

[11] Worldometers.info. Total coronavirus cases in india. https://www.worldometers.info/coronavirus/country/india/, 2021. Accessed on 6th May, 2021.

[12] Goverment of India. Covid19 statewise status. https://www.mygov.in/corona-data/covid19-statewise-status, 2020. Accessed on 6th May, 2021.

[13] Government of India. Covid-19 statewise status. https://www.mohfw.gov.in/, 2021. Accessed on 6th May, 2021.

[14] Office of the Registrar General & Census Commissioner, India. Provisional population totals. https://censusindia.gov.in/, 2011. Accessed on 6th May, 2021.

[15] Qifang Bi, Yongsheng Wu, Shujiang Mei, Chenfei Ye, Xuan Zou, Zhen Zhang, Xiaojian Liu, Lan Wei, Shaun A. Truelove, Tong Zhang, Wei Gao, Cong Cheng, Xiujuan Tang, Xiaoliang Wu, Yu Wu, Binbin Sun, Suli Huang, Yu Sun, Juncen Zhang, Ting Ma, Justin Lessler, and Tiejian Feng. Epidemiology and transmission of COVID-19 in 391 cases and 1286 of their close contacts in Shenzhen, China: a retrospective cohort study. The Lancet Infectious Diseases, 3099(20):1–9, 2020.

[16] Stephen A. Lauer, H. Grantz Kyra, Qifang Bi, Forrest K. Jones, Qulu Zheng, Hannah R. Meredith, Andrew S. Azman, Nicholas G. Reich, and Justin Lessler. The incubation period of coronavirus disease 2019 (covid-19) from publicly reported confirmed cases: Estimation and application. Annals of Internal Medicine, 172(9):577–582, 2020.

[17] Jacco Wallinga and Peter Teunis. Different epidemic curves for severe acute respiratory syndrome reveal similar impacts of control measures. American Journal of Epidemiology, 160(6):509–516, 2004.

[18] Tarun Bhatnagar, Sandip Mandal, Nimalan Arinaminpathy, Anup Agarwal, Amartya Chowdhury, Manoj Murhekar, RamanR Gangakhedkar, and Swarup Sarkar. Prudent public health intervention strategies to control the coronavirus disease 2019 transmission in india: A mathematical model-based approach. Indian Journal of Medical Research, 151(2):190, 2020.

[19] Emilia Vynnycky and Richard White. An Introduction to Infectious Disease Modelling. Oxford University Press, 2010.

[20] RM Anderson and MM Robert. Infectious Diseases of Humans. Oxford University Press, 1992.

[21] Derek Cummings and Justin Lessler. Infectious disease dynamics. http://www.interfetpthailand.net/file/June2007/Presentation/Dr.Derek/cummings.simulation.day.1.pdf, 2007.

[22] Luca Ferretti, Chris Wymant, Michelle Kendall, Lele Zhao, Anel Nurtay, Lucie Abeler-Dörner, Michael Parker, David Bonsall, and Christophe Fraser. Quantifying sars-cov-2 transmission suggests epidemic control with digital contact tracing. 368(6491), 2020.

[23] Joseph T. Wu, Kathy Leung, and Gabriel M. Leung. Nowcasting and forecasting the potential domestic and international spread of the 2019-nCoV outbreak originating in Wuhan, China: a modelling study. The Lancet, 395(10225):689–697, 2020.

[24] Kiesha Prem, Yang Liu, Timothy W Russell, Adam J Kucharski, Rosalind M Eggo, Nicholas Davies, Stefan Flasche, Samuel Clifford, Carl A B Pearson, James D Munday, Sam Abbott, Hamish Gibbs, Alicia Rosello, Billy J Quilty, Thibaut Jombart, Fiona Sun, Charlie Diamond, Amy Gimma, Kevin [van Zandvoort], Sebastian Funk, Christopher I Jarvis, W John Edmunds, Nikos I Bosse, Joel Hellewell, Mark Jit, and Petra Klepac. The effect of control strategies to reduce social mixing on outcomes of the covid-19 epidemic in wuhan, china: a modelling study. The Lancet Public Health, 5(5):e261 – e270, 2020.

[25] Qun Li, Xuhua Guan, Peng Wu, Xiaoye Wang, Lei Zhou, Yeqing Tong, Ruiqi Ren, Kathy S.M. Leung, Eric H.Y. Lau, Jessica Y. Wong, Xuesen Xing, Nijuan Xiang, Yang Wu, Chao Li, Qi Chen, Dan Li, Tian Liu, Jing Zhao, Man Liu, Wenxiao Tu, Chuding Chen, Lianmei Jin, Rui Yang, Qi Wang, Suhua Zhou, Rui Wang, Hui Liu, Yinbo Luo, Yuan Liu, Ge Shao, Huan Li, Zhongfa Tao, Yang Yang, Zhiqiang Deng, Boxi Liu, Zhitao Ma, Yanping Zhang, Guoqing Shi, Tommy T.Y. Lam, Joseph T. Wu, George F. Gao, Benjamin J. Cowling, Bo Yang, Gabriel M. Leung, and Zijian Feng. Early transmission dynamics in Wuhan, China, of novel coronavirus-infected pneumonia. New England Journal of Medicine, 382(13):1199–1207, 2020.

[26] J. A. Backer, D Klinkenberg, and J. Wallinga. Incubation period of 2019 novel coronavirus (2019-ncov) infections among travellers from wuhan, china, 20-28 january 2020. Euro Surveill., 25:2000062, Feb 2020.

[27] Saptarshi Chatterjee, Apurba Sarkar, Swarnajit Chatterjee, Mintu Karmakar, and Raja Paul. Studying the progress of COVID-19 outbreak in india using SIRD model. Indian Journal of Physics, jun 2020.

[28] Govt. of Indua. Ministry of health and family welfare. https://www.mohfw.gov.in/covid_vaccination/vaccination/faqs.html#what-to-expect-before-vaccination-2, Accessed on 6th May, 2021.

